# SIGNAL: A Scalable, Real-World Model for Rapid Intraoperative Molecular Classification of Gliomas Using Stimulated Raman Histology

**DOI:** 10.64898/2026.05.11.26350247

**Authors:** Nicolas K. Goff, John E. Markert, David Reinecke, Alexandra Springer, Anna M. Chen, Minjun Park, Gabrielle Malte, Katie Scotford-Broemmer, Sophia Hoonsbeen, Karen Eddy, Asadur Chowdury, Cheng Jiang, Akhil Kondepudi, Anna-Katharina Meissner, Gina Fürtjes, Nina Müller, Volker Neuschmelting, Melike Pekmezci, Jacob Young, Christian Freudinger, Matija Snuderl, Mitchel Berger, Shawn Hervey-Jumper, John G. Golfinos, Todd Hollon, Daniel A. Orringer

## Abstract

**Background:** Previous machine learning models to intraoperatively predict the molecular status of gliomas using stimulated Raman histology (SRH), such as DeepGlioma, have achieved high performance (91.5% accuracy) on curated datasets. However, when used intraoperatively, DeepGlioma (162M parameters) runs slowly on current SRH hardware and underperforms due to its lack of an image rejection mechanism and its validation on curated images. Here, we introduce SRH-Informed Glioma classificatioN with Attention Learning (SIGNAL) (27M parameters), a lighter model with a built-in attention-based rejection mechanism that outperforms DeepGlioma on uncurated clinical datasets.

**Methods:** SIGNAL was developed using 1.56 million SRH fields-of-view from 967 adult diffuse glioma patients collected between December 2017 and July 2025. We used 412 patients from NYU for training and internal validation and a multi-institutional, international cohort of 555 patients for testing. SIGNAL uses a ResNet50 backbone pretrained using a hierarchical contrastive loss function followed by a multi-head multi-layer perceptron (MLP). Using a patch-based attention threshold of 0.6, a final MLP was trained to predict glioma subtypes: glioblastoma, oligodendroglioma, or astrocytoma.

**Results:** SIGNAL outperformed DeepGlioma, achieving greater overall accuracy (90.10% vs. 72.59%) while running faster (16.0 vs. 6.7 patches/s). SIGNAL also outperformed DeepGlioma on all three molecular classification tasks, including IDH mutation (accuracy: 93.51% vs. 79.22%), 1p19q codeletion (93.51% vs. 88.31%), and ATRX loss (89.61% vs. 83.98%). SIGNAL’s attention mechanism had a strong positive linear correlation with mean patch cellularity (r=0.96, p<0.001) and a strong negative correlation with patch blood coverage (r=-0.99,p<0.001). Finally, subtype and molecular accuracy between tumor core and margin samples were equivalent despite significantly lower patch retention in tumor margins (44.5% vs 60.2%, p<0.0001).

**Conclusion:** SIGNAL is a lightweight model for intraoperative molecular classification of gliomas using SRH imaging. Its attention-based image quality filter allows for excellent performance, quick processing, and highly interpretable outputs critical for reliable use in intraoperative workflows.

**Brief 1-2 Sentence Description:** We present SIGNAL, a lightweight machine learning model for intraoperative molecular classification of diffuse gliomas using stimulated Raman histology, whose core innovation is a learned attention mechanism that filters diagnostically uninformative tissue, such as blood and acellular regions, before classification, enabling robust real-world generalizability. Validated on 555 patients across four international centers, SIGNAL outperforms the previous state-of-the-art model DeepGlioma on glioma subtype classification (90.10% vs. 72.59% accuracy) while running 2.4 times faster on intraoperative hardware.

## Background

The recent 2021 WHO CNS tumor classification change highlights how critical precision-based molecular diagnostics have become in glioma prognostication and management^1^. Despite the importance of these markers, such as IDH and 1p19q codeletion, in diagnosis and treatment of diffuse gliomas, clinical molecular testing by immunohistochemistry or DNA sequencing can take weeks, delaying patient care^2^. To mitigate these issues, Hollon, et. al. developed DeepGlioma, a rapid AI-based method of glioma molecular classification, using stimulated Raman histology (SRH) – a rapid, label-free optical imaging technique – for molecular classification of brain tumors. In this proof-of-concept study, DeepGlioma was able to achieve 91.5% accuracy for classification into the three major adult glioma subtypes: *IDH-*wildtype glioblastoma (GBM); *IDH*-mutant, 1p19q co-deleted oligodendroglioma; and *IDH*-mutant, *ATRX*-loss astrocytoma^3^.

Despite these promising results, DeepGlioma was developed on SRH images of high-quality specimens taken under optimal conditions from the tumor core. In practice, however, many samples are from margins that lack tumor tissue, are contaminated with hemorrhagic or cautery artifacts from surgical extraction or contain extensive necrotic regions not included in DeepGlioma’s learned patch distribution. Though DeepGlioma attempted to address this by filtering out samples with less than 10% tumor tissue, it generalizes poorly to uncurated datasets^4,5^. With heterogeneity within surgeons’ chosen sampling locations and sample tissues, there is a need for a robust algorithm with built-in filtering capable of rejecting low-quality samples.

Furthermore, while DeepGlioma can return a diagnosis faster than conventional frozen-section pathology, intraoperative workflows can take up to 15 minutes, depending on image size – well above published estimates. This gap reflects two compounding inefficiencies: image acquisition time for larger specimens, and the computational limitations of SRH hardware. Critically, DeepGlioma performs two sequential forward passes per patch – first through a ResNeXt-50 tumor classifier (~7.9 GFLOPs), then through a second ResNet-50-based transformer for molecular classification (~8.0 GFLOPs), meaning every tumor patch incurs nearly twice the compute of a single backbone pass. Though these limitations are secondary to the challenge of real-world generalizability, a more streamlined intraoperative workflow demands that they be addressed.

In this study, we present SRH-Informed Glioma classificatioN with Attention Learning (SIGNAL), a lighter model (27M parameters) with a built-in attention-based filter capable of rejecting diagnostically poor tissue and outperforming DeepGlioma on the largest SRH dataset of gliomas to date.

## Methods

This study was conducted following MI-CLAIM guidelines^6^.

### Patient Selection

Patients undergoing surgical resection or biopsy for suspected glioma at the University of Michigan (UM), New York University (NYU), the University of Cologne (UKK), and the University of California, San Francisco (UCSF) were imaged from December 2017 until July 2025. Pediatric patients (<18 years) and patients that did not have a final diagnosis of diffuse glioma were excluded from this study.

### Data Collection

Fresh tissue samples were collected intraoperatively from patients undergoing resection or biopsy for suspected glioma. Operating surgeons sampled grossly lesional-appearing tissue, which was then imaged using the NIO Laser Imaging System (Invenio Imaging, Inc.), previously described in detail^7,8^. After a technician selected a region of the tissue sample for SRH imaging, the specimen was stimulated using a 790nm pump beam and a Stokes beam tunable from 1,015-1,090nm. Two image channels were sequentially collected at Raman shifts of 2,845cm^-1^ (CH_2_; lipid) and 2,930cm^-1^ (CH_3_; protein/nucleic acid) as 1,000-pixel-wide strips. The first and last 50 pixels on the long axis of each strip were dropped to improve edge alignment^9^. Strips can then be concatenated and undergo recoloring for display as a virtual hematoxylin and eosin (H&E) - stained image or undergo further processing.

### Model Architecture

SIGNAL uses a ResNet50 backbone that outputs 2048-D feature vectors for each individual SRH patch. These features are passed to a common multi-layer perceptron (MLP) with three layers (512→256→128) and one attention head, followed by mutation-specific adapter and classifier layers (128→128→2)^10,11^. The attention head is then used as a patch quality filter (threshold = 0.6). A final MLP, trained on the output probabilities and filtered by attention, is then implemented to identify glioma subtypes. This MLP employs a four-layer architecture (3→128→128→64→3) with batch normalization and dropout (0.5, 0.4). This structure optimizes SIGNAL’s performance on both the molecular status and glioma subtype tasks.

**Figure 1:**
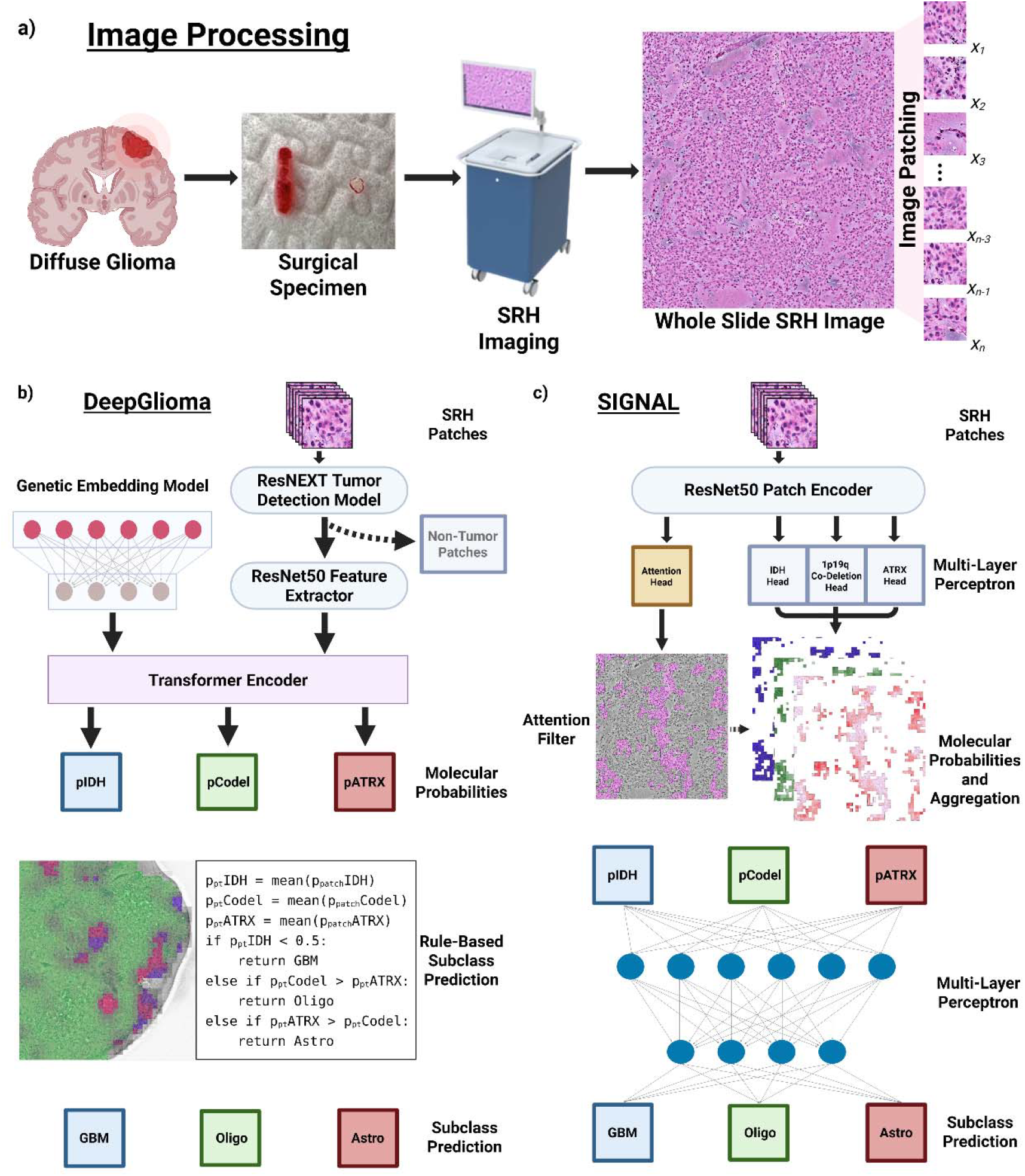
Comparison of SIGNAL and DeepGlioma Pipelines. a) The shared intraoperative workflow and imaging pipeline for both DeepGlioma and SIGNAL. Fresh surgical specimens from diffuse glioma patients are placed in a glass slide for SRH imaging. These images are then processed and split into 300×300 pixel patches. b) Flow diagram for DeepGlioma. Patches undergo two separate, sequential CNN forward passes – a ResNEXT for tumor segmentation followed by a ResNet50 for classification of tumor patches. These ResNet50 embeddings are passed with the outputs from the genetic embedding model into the transformer encoder, which outputs three molecular probabilities. These probabilities are used for rule-based subtype prediction. c) Flow diagram for SIGNAL. SRH patches are passed through a single ResNet50 backbone for feature extraction. The resultant embeddings are passed through the multi-head MLP, which includes three molecular heads and the attention head. Patches that pass the attention filter (threshold = 0.6) and their molecular head outputs are then aggregated to the patient level. Finally, these molecular probabilities are passed through the second MLP for subtype classification. CNN, convolutional neural network; MLP, multi-layer perceptron; SIGNAL, SRH-Informed Glioma classificatioN with Attention Learning; SRH, Stimulated Raman Histology.

**Figure 2:**
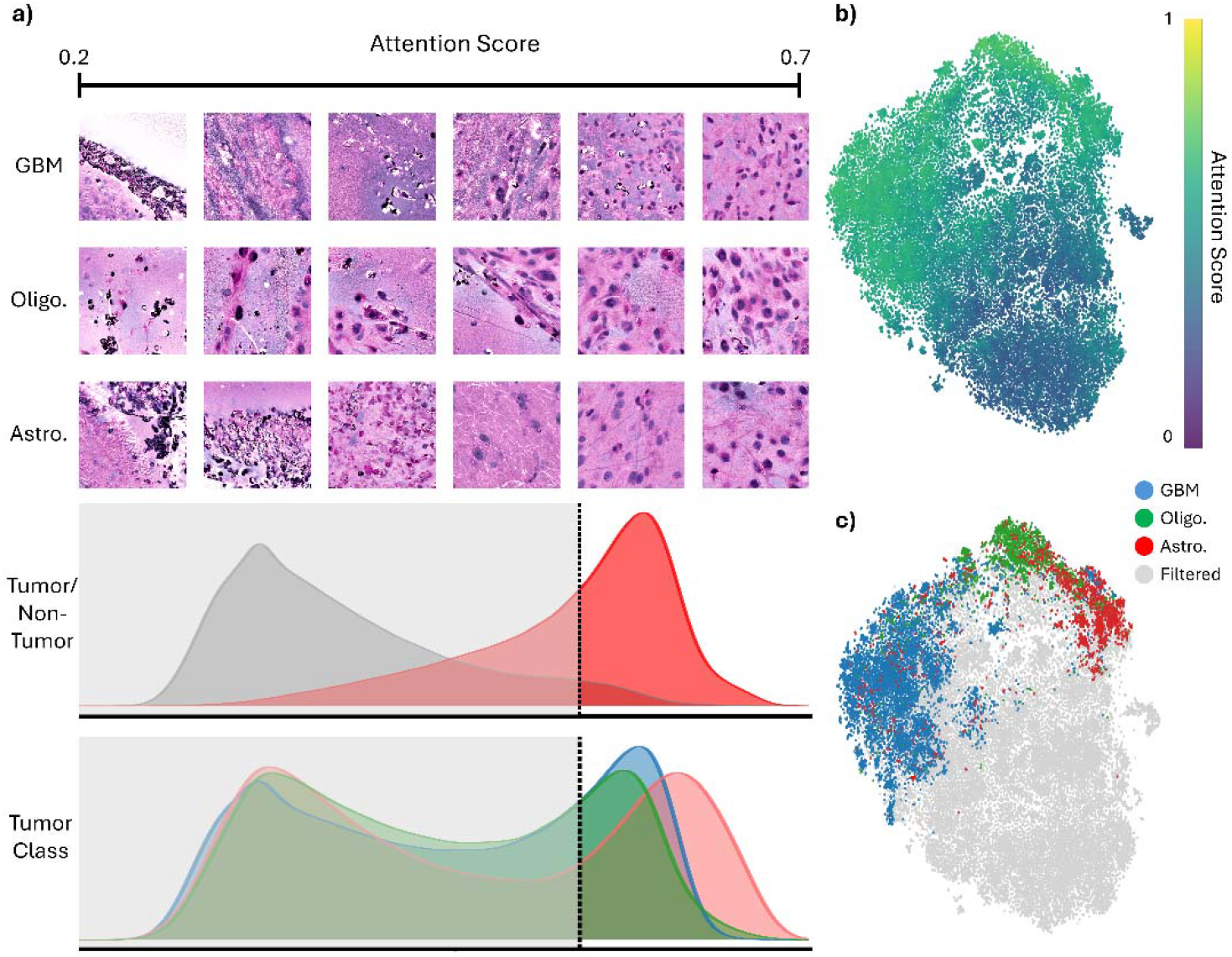
SIGNAL Attention Mechanism a) A series of example patches from each glioma subtype ordered by increasing attention score (top).The KDE plots show the distribution of attention scores when stratified by tumor/non-tumor status and subtype (bottom). The vertical dashed line denotes the attention filtering threshold of 0.6. The greyed-out area on the left side of the plot denotes patches that are filtered out by the attention mechanism. b) t-SNE plot for all patches colored by attention score. c) t-SNE plot for all patches colored by filtered status and glioma subtype. KDE, kernel density estimate; t-SNE, t-distributed Stochastic Neighbor Embedding.

**Figure 3:**
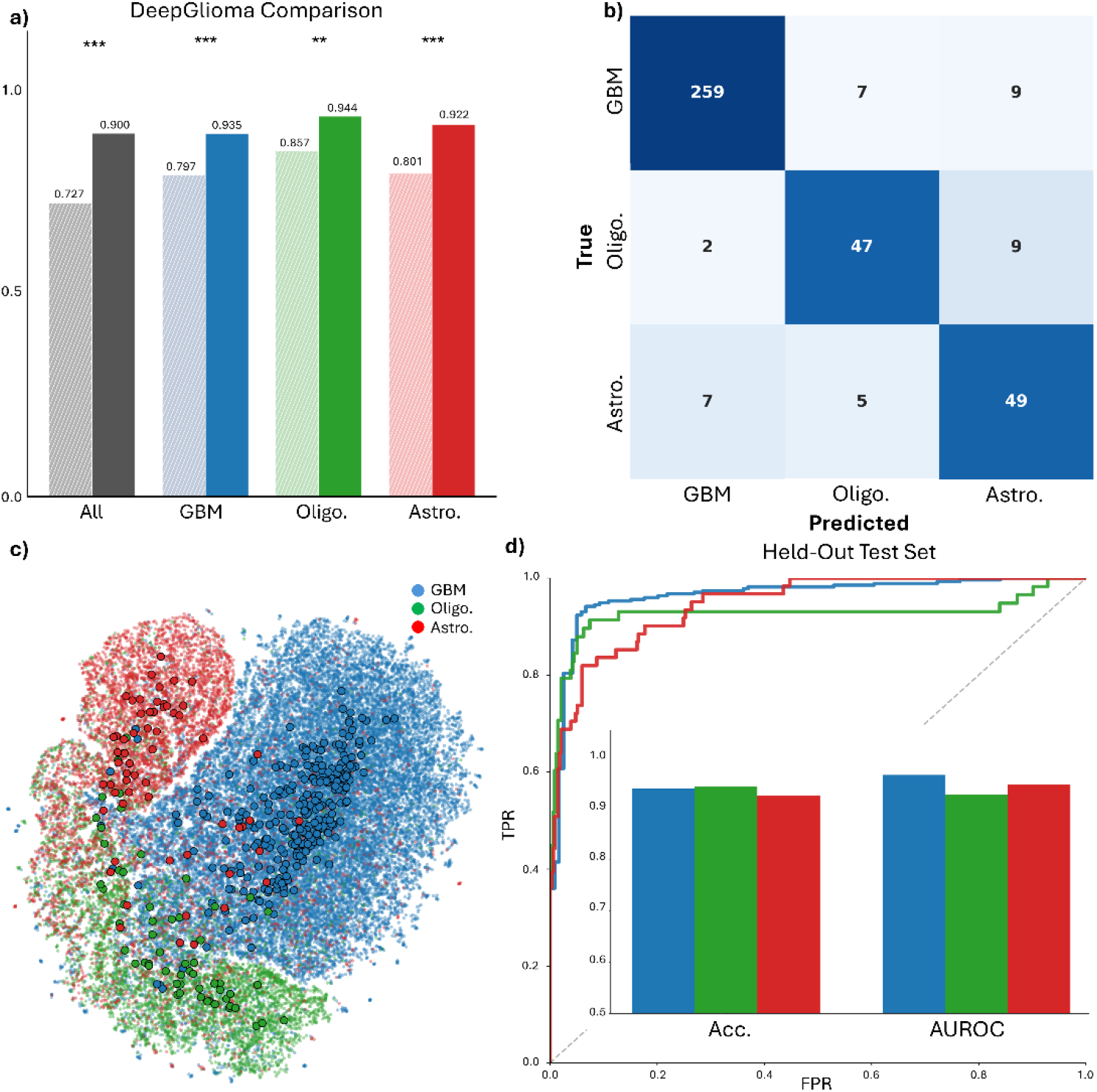
SIGNAL Diffuse Glioma Subtype Classification Results a) Bar plot comparing DeepGlioma (hatched bars) and SIGNAL’s (solid bars) overall and class accuracies on the UKK and UCSF test datasets. UM was excluded because of its use in training DeepGlioma. b) A confusion matrix by subtype for the full held-out test set (UCSF, UKK, UM). c) A t-SNE visualization of SRH representations from SIGNAL labelled by subtype. Small, translucent dots represent patches and large, solid dots represent patient mean patch locations. d) One-versus-rest ROC curves for glioblastoma (AUROC 0.965), oligodendroglioma (AUROC 0.855), and astrocytoma (AUROC 0.913); a bar graph showing accuracy and AUROC by subtype for the full held-out test set. AUROC, area under the receiver operating curve; SIGNAL, SRH-Informed Glioma classificatioN with Attention Learning; t-SNE, t-distributed Stochastic Neighbor Embedding; UCSF, the University of California San Francisco; UKK, the University of Cologne; UM, the University of Michigan. *, p<0.05; **, p<0.01; ***, p<0.001.

**Figure 4:**
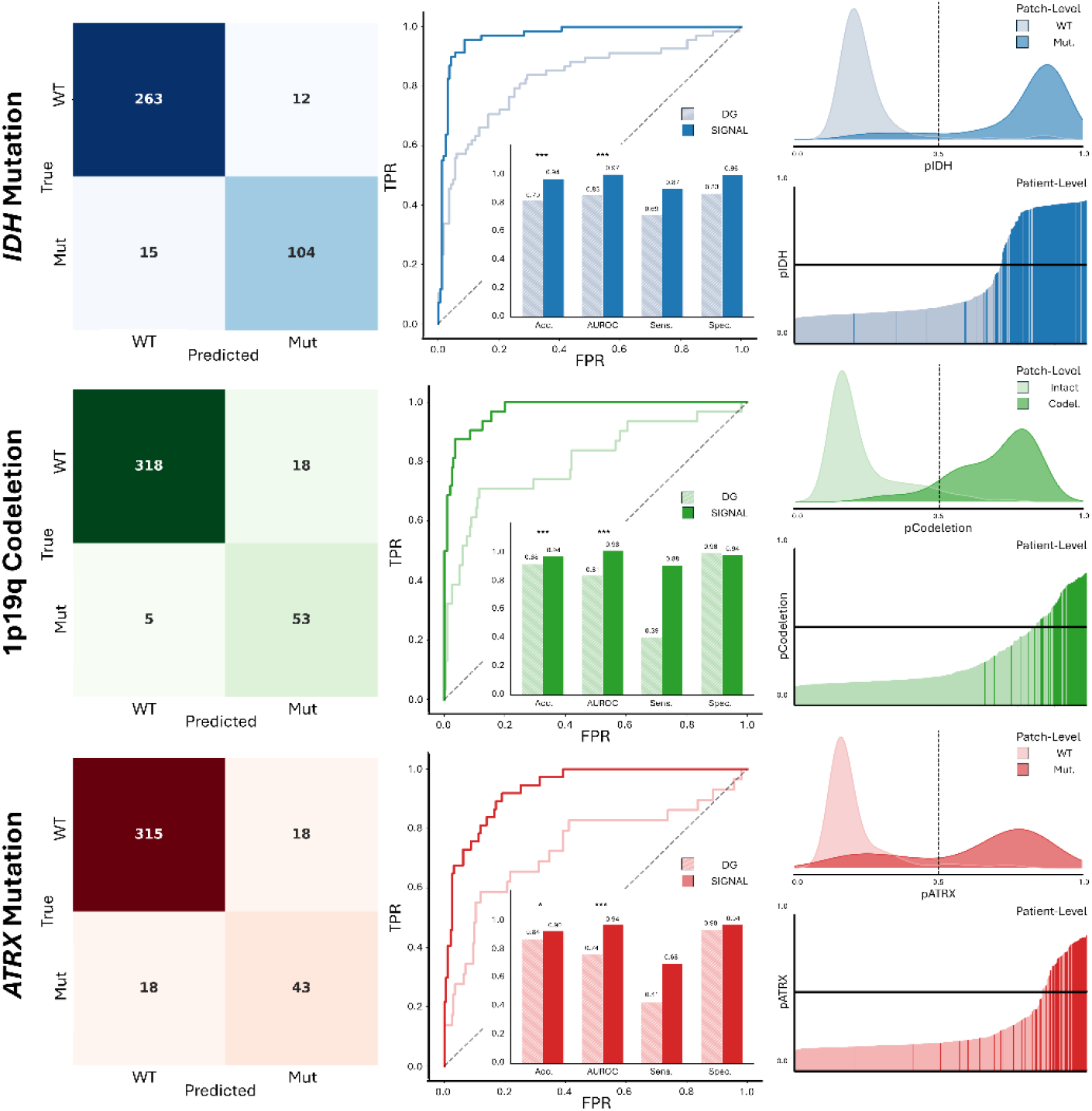
SIGNAL Molecular Classification Results. A grid demonstrating SIGNAL’s performance on the three molecular classification tasks – *IDH* mutation (AUROC 0.967), 1p19q codeletion (0.979), and *ATRX* loss (AUROC 0.926) (top to bottom). The left column contains a confusion matrix for each mutation on the full held-out test set (UCSF, UKK, UM). The center column is a comparison between DeepGlioma (light) and SIGNAL (dark) for each molecular task on the UCSF and UKK datasets, including ROC curves and a bar plot comparing accuracy, AUROC, sensitivity, and specificity. UM was excluded from this comparison due to its use in training DeepGlioma. The right column shows the distribution of patch-level molecular probabilities (top) and an ordered display of patient molecular predictions, colored by true molecular status (bottom). AUROC, area under the receiver operating curve; SIGNAL, SRH-Informed Glioma classificatioN with Attention Learning; UCSF, the University of California San Francisco; UKK, the University of Cologne; UM, the University of Michigan. *, p<0.05; **, p<0.01; ***, p<0.001.

**Figure 5:**
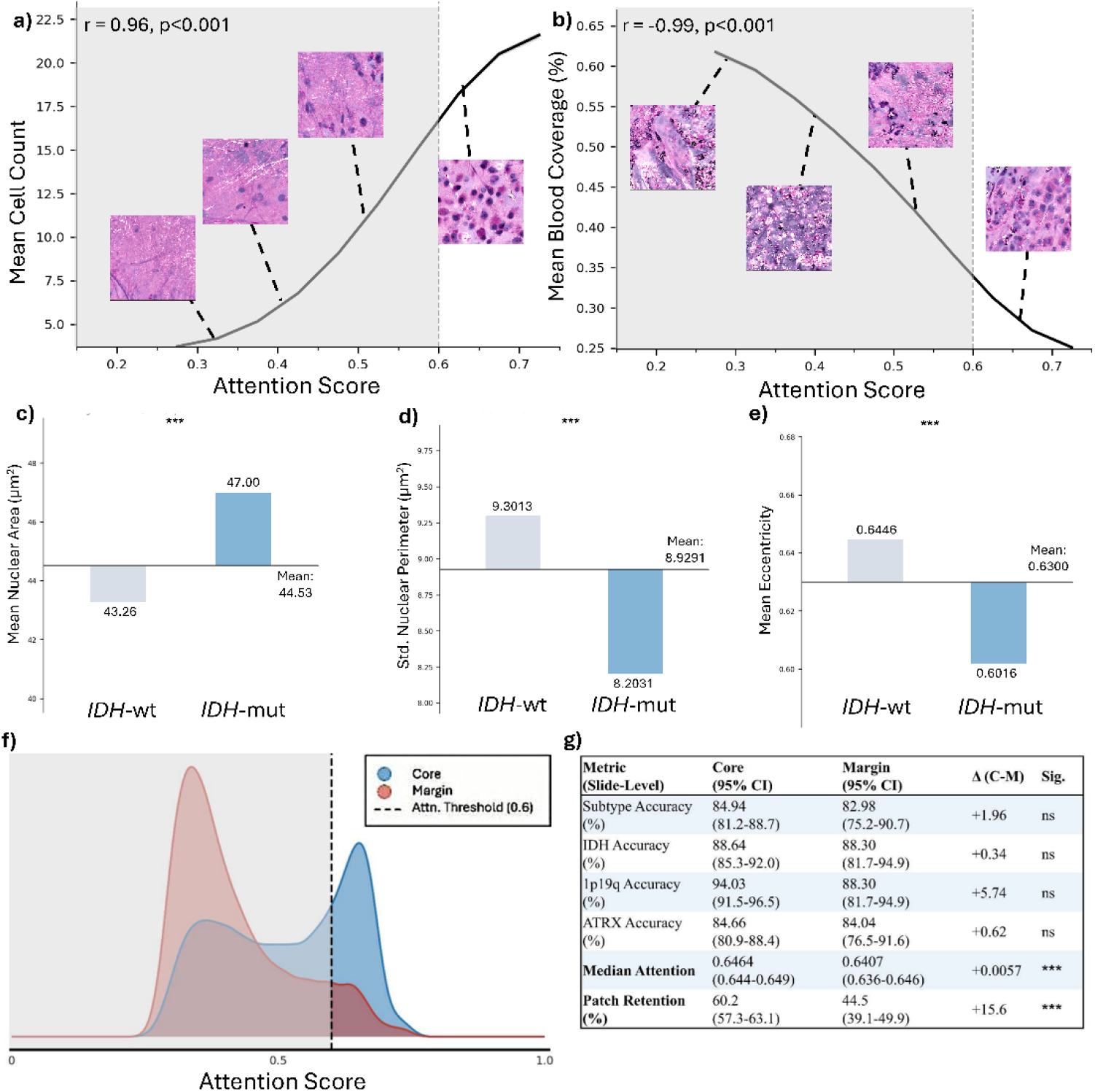
SIGNAL Interpretability Experiments and Clinical Deployment Considerations. a) A line plot demonstrating the relationship between mean patch cell count and SIGNAL attention score (r=0.96, p<0.001). Example patches with pseudo-H&E coloring show an increase in cellularity with increasing attention. Attention scores below the filtering threshold (0.6) are greyed out. b) A line plot showing the relationship between mean patch blood coverage and SIGNAL attention score (r=-0.99, p<0.001). Example patches with pseudo-H&E coloring show decreasing blood coverage with increasing attention. c-e) Bar plots comparing mean nuclear area (c), the standard deviation of nuclear perimeter, used as a proxy for nuclear pleomorphism (d), and mean nuclear eccentricity (e) for retained patches for *IDH*-wildtype and -mutant tumors. (f) A KDE plot of the attention distributions of samples taken from the tumor core (blue) and the tumor margin (red). The greyed-out region represents patches that were filtered out by SIGNAL’s attention mechanism. A table (g) showing the results of the Mann-Whitney U-Test between core and margin samples for subtype and mutation accuracy, median patch attention, and patch retention rates. H&E, hematoxylin and eosin; SIGNAL, SRH-Informed Glioma classificatioN with Attention Learning; CI, confidence interval; KDE, kernel density estimate. *, p<0.05; **, p<0.01; ***, p<0.001, ns, not significant.

### Training

Model training proceeded in three sequential stages. First, a ResNet50 backbone was pre-trained on all SRH patches from NYU using a weakly-supervised hierarchical contrastive loss optimized simultaneously for tumor/non-tumor discrimination and glioma subtype classification (GBM, oligodendroglioma, astrocytoma)^12^. Second, molecular classification heads (IDH mutation, 1p19q codeletion, ATRX mutation) were trained on features extracted from the frozen backbone using a multi-task learning framework with an integrated attention mechanism. Focal loss with class-specific weights addressed class imbalance across underrepresented markers. Attention scores were used to filter non-informative patches, with the optimal threshold selected via validation set sweep; patients with <10% of patches retained were also filtered. Finally, an MLP integrating the three molecular predictions into a unified WHO classification was trained on attention-filtered patches, followed by fine-tuning on slide-level aggregated predictions weighted by attention score. Full training hyperparameters are provided in Supplementary Methods.

### Evaluation

The model was evaluated on a multi-institutional, held-out test set. Metrics, including accuracy and mean area under the receiver operating curve (AUROC) were calculated for each molecular task and the subtype task. Attention metrics for tumor and non-tumor patches, as classified by a model previously published by our group, and for each individual class were quantified and compared using the Wilcoxon rank-sum test^4^.

To compare SIGNAL with DeepGlioma, we evaluated both models on test datasets from UKK and UCSF on the molecular and subtype tasks. UM cases were excluded from this comparison due to their use in training DeepGlioma to avoid any possibility of data leakage or institutional confounding. McNemar’s test was used for paired binary molecular comparisons, the bootstrap test (n_iter_=1,000) for multi-class subtype accuracy, and De Long’s test for AUROC comparisons between correlated classifiers. All statistical tests were conducted using a significance level of α=0.05.

### Interpretability

To verify the validity of our attention filter, we used a previously developed SRH cell detection algorithm trained to detect macrophages, red blood cells, and cell nuclei from SRH patches^13^. From this algorithm, we extracted morphometric measurements including nuclear area, perimeter, and eccentricity, as well as nuclear count and red blood cell patch coverage. Welch’s two-sample t-test was used to compare patient means between IDH-wildtype and IDH mutant tumors. Simple linear regression was then performed to evaluate the relationship between attention (binned at 0.05 intervals from 0 to 1) and average red blood cell coverage and nuclear count.

### Tissue Requirement and Robustness Analysis

To identify minimum tissue requirements, we investigated the relationship between retained patch count and model performance using point-biserial correlation and compared retention rates across tumor types. To characterize model behavior across varying tissue availability, we performed progressive patch subsampling (k = 5 to 250 patches, 1000 iterations per k) on the multi-institutional test set, measuring accuracy and prediction stability at each k. Normal tissue rejection was tested by running inference on 38 images from 19 cadaveric brain samples and calculating overall tissue retention and attention statistics. Similarly, slide-level performance and attention statistics were calculated for samples taken from the tumor core and tumor margin and compared using the Mann-Whitney U-test.

## Results

### Patient Cohort

From October 2019 until July 2025, we identified 412 patients at our institution that underwent either resection or biopsy for glioma that were imaged with our SRH system. 66.7% of these patients’ tumors were glioblastomas, 13.3% oligodendrogliomas, and 19.9% astrocytomas. They were then split at the patient level into training and validation sets, with 333 patients and 797,490 fields-of-view (FOVs) used for training and 79 patients with 187,290 FOVs used for validation.

The model was then externally validated using an international, multi-institutional cohort sourced from UM, UCSF, and UKK. In total, 555 patients with 575,928 FOVs were included in our test set, with 69.0% glioblastoma, 13.3% oligodendroglioma, and 17.7% astrocytoma.

### Attention Mechanism

The mean tumor patch attention score was 0.594±0.08, while the mean non-tumor patch attention score was 0.408±0.09 (z=543.7, p<0.0001). Mean attention scores for tumor patches from each class were 0.590, 0.627, and 0.580 for glioblastoma, oligodendroglioma, and astrocytoma, respectively.

### Subtype Classification

The primary endpoint for our study was model performance on the glioma subtype classification task. Using an attention threshold of 0.6, our model achieved an overall accuracy of 90.10% on the external test set, with class AUROCs of 0.965, 0.855, and 0.913 for GBM, oligodendroglioma, and astrocytoma, respectively, with a macro-averaged F1 of 0.841 and weighted F1 of 0.903.

SIGNAL outperformed DeepGlioma on our dataset of 430 patients from UCSF and UKK, achieving an overall accuracy of 90.10%, as compared to DeepGlioma’s 72.59%. Similarly, SIGNAL demonstrated better class accuracies, reaching 93.5% vs 85.7% for GBMs, 94.4% vs 86.9% for oligodendrogliomas, and 92.2% vs 86.9% for astrocytomas. The bootstrap test showed that all differences were statistically significant.

### Molecular Classification

The secondary endpoints for this study were model performance on the three major molecular targets involved in adult glioma classification, according to the 2021 WHO guidelines – *IDH*, 1p19q codeletion, and *ATRX* loss. Here too, SIGNAL achieved state-of-the-art performance, reaching 93.51% accuracy in predicting *IDH* status on the held-out test set (DeepGlioma: 79.22%, p<0.0001). These gains in performance were also seen for the 1p19q codeletion and *ATRX* tasks – with SIGNAL achieving 93.51% accuracy (DeepGlioma: 88.31%, p<0.0001) and 89.61% accuracy (DeepGlioma: 83.98%, p<0.0001), respectively.

### Interpretability

Patch cell count showed a strong positive linear correlation with attention (r=0.96, p<0.001), while red blood cell coverage showed a strong negative correlation with attention (r=-0.99, p<0.001). Furthermore, among high-attention patches, we found that the standard deviation of nuclear perimeter (9.30 vs. 8.20µm, p<0.0001) and mean nuclear eccentricity (0.6446 vs. 0.6016, p<0.0001) were higher among *IDH* wildtype tumor patches, consistent with greater nuclear pleomorphism found in glioblastoma. Macrophage counts were also increased in high-attention *IDH* wild-type patches (2.47 vs 2.06 macrophages/patch, p<0.0001). Interestingly, mean nuclear area was lower among *IDH-*wildtype tumors compared to *IDH-*mutants (43.26 vs 47.00 µm^2^, p<0.0001), consistent with previous SRH morphometric findings^14^. These results demonstrate that SIGNAL’s attention mechanism learned real, human-interpretable image features that align with known histopathological features of the various subtypes of glioma.

### Clinical Deployment Considerations

We first compared SIGNAL’s performance on a subset of images from the tumor core (n=628) and the tumor margin (n=511) in our test set. SIGNAL performed similarly between the two groups for the subtype task and all three molecular tasks, with no statistically significant differences in accuracy. There were, however, significant differences in median attention (0.6464 vs 0.6407, p=0.0065) and patch retention rates (60.2% vs 44.5%, p<0.0001).

We then compared the computational efficiency of SIGNAL and DeepGlioma under conditions mirroring intraoperative deployment: 500 randomly sampled mosaics processed on four CPU cores. Memory footprint was statistically identical between models (Welch’s t-test=0.0001, p=0.9999). SIGNAL achieved a mean throughput of 16.0 patches/second versus 6.7 patches/second for DeepGlioma (Mann-Whitney U=0.0, p<0.0001) – a 2.4-fold improvement. This efficiency gain is explained by fundamental architectural differences in compute allocation. SIGNAL processes every patch through a single ResNet-50 backbone (7.67 GFLOPs), with the downstream attention filter and MLP classifier contributing less than 0.1% of total compute. In contrast, DeepGlioma’s cost per patch scales with tissue composition: non-tumor patches incur ~7.9 GFLOPs, while tumor patches – which trigger the molecular classifier – incur ~15.9 GFLOPs as a second full ResNet-50 pass is performed inside the transformer. This variable compute burden explains SIGNAL’s near-perfect throughput scaling with mosaic size (R^2^=0.9999) compared to DeepGlioma’s substantially lower linearity (R^2^=0.8690): as tumor fraction varies across patients, so does DeepGlioma’s per-patient cost.

Next, we investigated the amount of tissue required by SIGNAL for stable and accurate prediction. We found that there was a weak positive correlation (r=0.118, p=0.0061) between the number of retained patches for a patient and prediction correctness. The threshold to reach 99.5% prediction stability was 175 patches. With SIGNAL’s patch retention rate of 30%, two 2.5×2.8mm slides are needed to reach this threshold, on average – leading to an estimated intraoperative run time of under 10 minutes. To assess whether excluded patients represented genuine diagnostic failures, we compared model performance at threshold=0.40 – the highest threshold retaining all patients – between those who subsequently passed or failed the threshold=0.60 filter. Patients retained at threshold=0.60 achieved 81% accuracy at threshold=0.40, compared to 35% among those excluded, suggesting the attention mechanism functions as a confidence-based triage tool that selectively rejects cases with insufficient diagnostic signal rather than arbitrarily reducing cohort size.

Finally, we ran SIGNAL on a series of 38 images sampled from 19 different brain regions of healthy cadaveric brain tissue. The attention mechanism successfully rejected all 38 images as non-tumor. The patch attention range for this dataset was 0.2752-0.6121. Only 7 images had patches with attention scores over the 0.6 threshold, sampled from the putamen, sensory cortex, hippocampus, thalamus, globus pallidus internus, and globus pallidus externus. The maximum tissue retention rate was 0.18% (globus pallidus externus), demonstrating SIGNAL’s ability to filter out normal tissue prior to classification.

## Discussion

Despite the recognition of the importance of IDH, 1p19q codeletion, and ATRX loss in diagnosing diffuse gliomas, the current standard of care does not allow for their detection in a timely manner. This information is important for diagnosis, prognostication, and may also provide context for surgical decision-making. Among IDH-mutant tumors, extent of resection has been shown to have differential prognostic impact across subtypes, though the implications of these differences for intraoperative management are under debate, and the current standard of care favors maximal safe resection regardless of subtype^15–18^. Here, we present SIGNAL, a lightweight glioma classifier achieving 90.10% subtype accuracy on a multi-institutional, uncurated dataset versus DeepGlioma’s 72.59%, which we attribute to broader training and a learned attention-based quality filter.

There are several issues we found with DeepGlioma that we sought to improve with SIGNAL. First and foremost is DeepGlioma’s limited robustness when dealing with real-world data. Because DeepGlioma was trained on a dataset of highly representative images from a select group of cases and came with a very weak rejection mechanism (<10% tumor tissue), it has no built-in failure mode^3^. This leads to the DeepGlioma generating predictions for low-quality tissue, significantly hindering its performance without a human in the loop – a common problem for medical diagnostic models^19,20^. SIGNAL addresses these issues by combining an unfiltered training set with a learned attention mechanism, allowing it to recognize a wider range of tissue quality while automatically filtering uninformative patches, similar to previous models that used instance-level attention as a quality filter^10,21,22^. Importantly, though margin tissue had a much lower rate of retention compared to core tissue (44.5% vs 60.2%), classification accuracy was statistically equivalent across all tasks, demonstrating that SIGNAL’s attention mechanism can compensate for suboptimal sampling locations. However, when possible, sampling from grossly lesional tissue is recommended to avoid low tissue retention rates.

A second key improvement in SIGNAL is architectural consolidation. DeepGlioma’s pipeline requires two independent forward passes per patch, doubling the computational load on tumor patches to ~15.9 GFLOPs.^3^. SIGNAL’s single ResNet-50 backbone handles feature extraction for both quality filtering and molecular classification, with the downstream MLP contributing negligible additional compute. The result is a model that is 2.4x faster than DeepGlioma, and whose runtime scales linearly and predictably with mosaic size^23,24^.

Finally, the correlation of SIGNAL’s attention scores with known, interpretable markers such as cell count and blood coverage represents a significant leap forward from previous “black box” models. Demonstrating the relationship between these features and attention scores allows for increased transparency in model decision-making which is essential in medical applications of artificial intelligence^25^.

### Limitations

When SIGNAL’s attention filter rejects a sample, the appropriate clinical response is re-imaging an adjacent region rather than abandoning the specimen – analogous to an indeterminate intraoperative frozen section prompting additional sampling. Since SRH is non-destructive, this adds imaging time but requires no further tissue sacrifice, which is particularly relevant in eloquent cortex or biopsy-only cases. The filter reduced evaluable patients from 555 to 394 (~29%); this exclusion rate reflects genuine diagnostic uncertainty rather than arbitrary tissue loss, as 65% of excluded patients would have been misclassified at the most lenient viable threshold, compared to SIGNAL’s 90.10% accuracy.

A second limitation of this study is label quality. Patch-level tumor/non-tumor labels were derived from DeepGlioma’s segmentation model, introducing a risk that SIGNAL’s attention mechanism partially inherited its biases. Furthermore, molecular and subtype labels were weak, patient-level assignments rather than patch-level ground truth; this is a common limitation across whole-slide imaging models, not only with SRH models like DeepGlioma, but also in other computational pathology settings^26,27^. Together, these sources of label noise likely represent the primary ceiling on model performance and motivate future work incorporating manually annotated patch-level labels.

Finally, SIGNAL was developed and validated exclusively on adult diffuse gliomas per the 2021 WHO classification – *IDH*-wildtype GBM; *IDH*-mutant, 1p19q-codeleted oligodendroglioma; and *IDH*-mutant, 1p19q-intact astrocytoma. Other tumors, such as lower-grade glioneuronal tumors, BRAF-driven gliomas, and pediatric entities were not included in this study and SIGNAL’s outputs on these tumor types may be inaccurate. SRH images that raise suspicion for one of these tumors should be reviewed by a neuropathologist for accurate classification.

### Future Directions

Adding more molecular heads to SIGNAL, such as *CDKN2A/B, MGMT* methylation, or BRAF, may prove useful in both intraoperative characterization and prognostication of gliomas^28–30^. Additional molecular heads would both enrich the feature representations learned by the shared backbone and increase the dimensionality of inputs to the final classification MLP, potentially improving subtype separation and reducing sensitivity to errors in any single predictive dimension^31–33^. Furthermore, SIGNAL’s application to non-glioma CNS tumors warrants investigation. While the attention mechanism’s learned rejection of normal brain suggests potential generalizability, whether it extends to non-glioma tumor tissue – such as meningioma or metastasis – remains untested. Further developing the model to perform either of these tasks would increase its clinical utility.

## Data Availability

All data produced in the present study are available upon reasonable request to the authors.

## Conclusion

SRH is a powerful tool for rapid, non-destructive, label-free intraoperative classification of gliomas. However, previous models, such as DeepGlioma, were validated on highly curated datasets and utilized inefficient model architectures^4^. SIGNAL relies on a built-in quality filter that allows it to outperform DeepGlioma in both molecular and subtype classification tasks on real-world, complex datasets while improving inference speed and model interpretability.

## References

1. Louis DN, Perry A, Wesseling P, et al. The 2021 WHO Classification of Tumors of the Central Nervous System: a summary. Neuro Oncol 2021;23(8):1231–1251. (In eng). DOI: 10.1093/neuonc/noab106.

2. Penkova A, Kuziakova O, Gulaia V, et al. Comprehensive clinical assays for molecular diagnostics of gliomas: the current state and future prospects. Frontiers in Molecular Biosciences 2023;Volume 10 - 2023 (Review) (In English). DOI: 10.3389/fmolb.2023.1216102.

3. Hollon T, Jiang C, Chowdury A, et al. Artificial-intelligence-based molecular classification of diffuse gliomas using rapid, label-free optical imaging. Nature Medicine 2023;29(4):828–832. DOI: 10.1038/s41591-023-02252-4.

4. Hollon TC, Pandian B, Adapa AR, et al. Near real-time intraoperative brain tumor diagnosis using stimulated Raman histology and deep neural networks. Nature Medicine 2020;26(1):52–58. DOI: 10.1038/s41591-019-0715-9.

5. Hollon TC, Pandian B, Urias E, et al. Rapid, label-free detection of diffuse glioma recurrence using intraoperative stimulated Raman histology and deep neural networks. Neuro-Oncology 2021;23(1):144–155. DOI: 10.1093/neuonc/noaa162.

6. Norgeot B, Quer G, Beaulieu-Jones BK, et al. Minimum information about clinical artificial intelligence modeling: the MI-CLAIM checklist. Nature Medicine 2020;26(9):1320–1324. DOI: 10.1038/s41591-020-1041-y.

7. Freudiger CW, Min W, Saar BG, et al. Label-Free Biomedical Imaging with High Sensitivity by Stimulated Raman Scattering Microscopy. Science 2008;322(5909):1857–1861. DOI: 10.1126/science.1165758.

8. Freudiger CW, Yang W, Holtom GR, Peyghambarian N, Xie XS, Kieu KQ. Stimulated Raman scattering microscopy with a robust fibre laser source. Nature Photonics 2014;8(2):153–159. DOI: 10.1038/nphoton.2013.360.

9. Orringer DA, Pandian B, Niknafs YS, et al. Rapid intraoperative histology of unprocessed surgical specimens via fibre-laser-based stimulated Raman scattering microscopy. Nature Biomedical Engineering 2017;1(2):0027. DOI: 10.1038/s41551-016-0027.

10. Vaswani A, Shazeer N, Parmar N, et al. Attention is All you Need. In: Guyon I, Luxburg UV, Bengio S, et al., eds. 2017.

11. Hernández A, Amigó JM. Attention Mechanisms and Their Applications to Complex Systems. Entropy (Basel) 2021;23(3) (In eng). DOI: 10.3390/e23030283.

12. Jiang C, Hou X, Kondepudi A, et al. Hierarchical discriminative learning improves visual representations of biomedical microscopy. 2023.

13. Bhattacharya A, Alber D, Hollon T. Single-cell phenotyping using optical imaging and artificial intelligence. Machine Learning for Healthcare 2022. Durham, NC 2022.

14. Reinecke D, Goff N, Markert J, et al. P04.07.A AI-BASED NUCLEAR MORPHOMETRY REVEALS DISTINCT MORPHOLOGICAL PATTERNS BETWEEN IDH-MUTANT, IDH-WILDTYPE, AND NON-TUMOROUS CELLS IN STIMULATED RAMAN HISTOLOGY. Neuro-Oncology 2025;27(Supplement_3):iii60–iii60. DOI: 10.1093/neuonc/noaf193.193.

15. van der Vaart T, Wijnenga MMJ, van Garderen K, et al. Differences in the Prognostic Role of Age, Extent of Resection, and Tumor Grade between Astrocytoma IDHmt and Oligodendroglioma: A Single-Center Cohort Study. Clin Cancer Res 2024;30(17):3837–3844. (In eng). DOI: 10.1158/1078-0432.Ccr-24-0901.

16. Ding X, Wang Z, Chen D, et al. The prognostic value of maximal surgical resection is attenuated in oligodendroglioma subgroups of adult diffuse glioma: a multicenter retrospective study. J Neurooncol 2018;140(3):591–603. (In eng). DOI: 10.1007/s11060-018-2985-3.

17. Hervey-Jumper SL, Zhang Y, Phillips JJ, et al. Interactive Effects of Molecular, Therapeutic, and Patient Factors on Outcome of Diffuse Low-Grade Glioma. Journal of Clinical Oncology 2023;41(11):2029–2042. DOI: 10.1200/JCO.21.02929.

18. Chen JS, Oh JY, Hollon TC, Hervey-Jumper SL, Young JS, Berger MS. The Intraoperative Utility of Raman Spectroscopy for Neurosurgical Oncology. Cancers (Basel) 2025;17(24) (In eng). DOI: 10.3390/cancers17243920.

19. Wu E, Wu K, Daneshjou R, Ouyang D, Ho DE, Zou J. How medical AI devices are evaluated: limitations and recommendations from an analysis of FDA approvals. Nature Medicine 2021;27(4):582–584. DOI: 10.1038/s41591-021-01312-x.

20. Goetz L, Seedat N, Vandersluis R, van der Schaar M. Generalization—a key challenge for responsible AI in patient-facing clinical applications. npj Digital Medicine 2024;7(1):126. DOI: 10.1038/s41746-024-01127-3.

21. Ren M, Zeng W, Yang B, Urtasun R. Learning to Reweight Examples for Robust Deep Learning. In: Jennifer D, Andreas K, eds. Proceedings of the 35th International Conference on Machine Learning. Proceedings of Machine Learning Research: PMLR; 2018:4334--4343.

22. Jiang L, Zhou Z, Leung T, Li L-J, Fei-Fei L. MentorNet: Learning Data-Driven Curriculum for Very Deep Neural Networks on Corrupted Labels. In: Jennifer D, Andreas K, eds. Proceedings of the 35th International Conference on Machine Learning. Proceedings of Machine Learning Research: PMLR; 2018:2304--2313.

23. Lema DG, Usamentiaga R, García DF. Quantitative comparison and performance evaluation of deep learning-based object detection models on edge computing devices. Integration 2024;95:102127. DOI: 10.1016/j.vlsi.2023.102127.

24. Mattern A, Gerdes H, Grunert D, Schmitt RH. A comparison of transformer and CNN-based object detection models for surface defects on Li-Ion Battery Electrodes. Journal of Energy Storage 2025;105:114378. DOI: 10.1016/j.est.2024.114378.

25. Kondepudi A, Pekmezci M, Hou X, et al. Foundation models for fast, label-free detection of glioma infiltration. Nature 2025;637(8045):439–445. DOI: 10.1038/s41586-024-08169-3.

26. Chen RJ, Ding T, Lu MY, et al. Towards a general-purpose foundation model for computational pathology. Nature Medicine 2024;30(3):850–862. DOI: 10.1038/s41591-024-02857-3.

27. Hoang DT, Shulman ED, Turakulov R, et al. Prediction of DNA methylation-based tumor types from histopathology in central nervous system tumors with deep learning. Nat Med 2024;30(7):1952–1961. (In eng). DOI: 10.1038/s41591-024-02995-8.

28. Lu VM, O’Connor KP, Shah AH, et al. The prognostic significance of CDKN2A homozygous deletion in IDH-mutant lower-grade glioma and glioblastoma: a systematic review of the contemporary literature. Journal of Neuro-Oncology 2020;148(2):221–229. DOI: 10.1007/s11060-020-03528-2.

29. Schreck KC, Langat P, Bhave VM, et al. Integrated molecular and clinical analysis of BRAF-mutant glioma in adults. npj Precision Oncology 2023;7(1):23. DOI: 10.1038/s41698-023-00359-y.

30. Hegi Monika E, Diserens A-C, Gorlia T, et al. MGMT Gene Silencing and Benefit from Temozolomide in Glioblastoma. New England Journal of Medicine 2005;352(10):997–1003. DOI: 10.1056/NEJMoa043331.

31. Bengio Y, Courville A, Vincent P. Representation Learning: A Review and New Perspectives. IEEE Transactions on Pattern Analysis and Machine Intelligence 2013;35(8):1798–1828. DOI: 10.1109/TPAMI.2013.50.

32. Dietterich TG. Ensemble Methods in Machine Learning. Multiple Classifier Systems. Berlin, Heidelberg: Springer Berlin Heidelberg; 2000:1–15.

33. Zhang Y, Yang Q. A Survey on Multi-Task Learning. IEEE Transactions on Knowledge and Data Engineering 2022;34(12):5586–5609. DOI: 10.1109/TKDE.2021.3070203.

