## Supplemental Appendix for "SIGNAL: A Scalable, Real-World Model for Rapid Intraoperative Molecular Classification of Gliomas Using Stimulated Raman Histology"

**Supplementary Appendix**

### Supplementary Methods

*Pre-Processing*

Model training and inference were done using raw SRH images rather than virtual H&E-stained images. 900 pixel-wide strips first underwent field flattening correction and Fourier-transform-based co-registration to account for variations in pixel intensity and natural movements in tissue during imaging, respectively. Images were divided into 300 x 300-pixel patches without overlap. A third channel was generated by performing pixel-wise subtraction between the CH_3_ and CH_2_ channels to create RGB images (R: CH_3_ - CH_2_, G: CH_2_, B: CH_3_). Channel intensity values were normalized to (0,1).

*Training Hyperparameters*

All models were trained using the AdamW optimizer. The ResNet50 backbone was pre-trained for up to 100,000 steps with an initial learning rate of 1×10⁻², a 10,000-step linear warmup followed by cosine annealing, and early stopping. The augmentations used to create positive pairs at the image level of the hierarchical loss function include flipping, Gaussian noise, color jitter, autocontrast, solarization, sharpness adjustment, affine transformation, and random erasing. Molecular classification heads were trained for 13 epochs (lr=1×10⁻⁵, early stopping patience=10), incorporating uncertainty regularization and attention diversity constraints. The final subtype classification MLP was trained for 8 epochs (lr=5×10⁻⁷, early stopping patience=30). Focal loss class-specific weights were as follows: αIDH=[1.0,2.0], αCodel=[1.0,2.0], αATRX=[1.0,2.5], γ=2.0 across all molecular heads; the subtype MLP used weighted focal loss (γ=2.0, α=[1.0,1.5,2.0]) scaled by molecular prediction confidence.

*Tissue Requirement and Robustness Analysis*

Retention rate across tumor types (GBM, oligodendroglioma, astrocytoma) was compared using the Kruskal-Wallis test. The relationship between the number of retained patches and prediction correctness was quantified using point-biserial correlation.

For progressive patch subsampling, k patches were randomly sampled per patient (k = 5 to 250 in increments of 5) and predictions were generated using SIGNAL. Each sampling was repeated 1000 times to quantify variability. Prediction stability was defined as the agreement rate between k-patch predictions and full-tissue predictions (using all available patches). Accuracy at each k was computed against ground truth labels. Mean and standard error were calculated across sampling iterations for both metrics.

Cadaveric brain samples were obtained from 19 regions, including the sensory, motor, and visual cortices, the limbic system, putamen, thalamus, globus pallidus internus and externus, hypothalamus, pineal gland, pons, and cerebellum; samples of the olfactory and optic nerves were also included. Two images were acquired per region for a total of 38 images. Overall tissue retention, patch attention range, and the percentage of area covered by unfiltered patches were calculated for each image.

| Table S1. Training Dataset Demographic Summary | | | | | | |
| --- | --- | --- | --- | --- | --- | --- |
| Characteristic | **Overall** | **GBM** | | **Oligo** | | **Astro** |
| Demographics |  |  |  | |  | |
| N | 412 | 275 | 55 | | 82 | |
| Age (years), median [IQR] | 56.8 (42.5-67.0) | 61.8 (52.9-72.1) | 39.1 (34.1-49.8) | | 40.5 (31.2-48.1) | |
| < 35 | 62 (15.0%) | 16 (5.8%) | 15 (27.3%) | | 31 (37.8%) | |
| 35-65 | 221 (53.6%) | 138 (50.2%) | 37 (67.3%) | | 46 (56.1%) | |
| > 65 | 127 (30.8%) | 120 (43.6%) | 3 (5.5%) | | 4 (4.9%) | |
| Unknown | 2 (0.4%) | 1 (0.0%) | 0 (0.0%) | | 1 (1.2%) | |
| Sex |  |  |  | |  | |
| Female | 161 (39.1%) | 102 (37.1%) | 26 (47.3%) | | 33 (40.2%) | |
| Male | 242 (58.7%) | 170 (61.8%) | 26 (47.3%) | | 46 (56.1%) | |
| Race |  |  |  | |  | |
| White not of Hispanic American Origin | 275 (66.7%) | 191 (69.5%) | 37 (67.3%) | | 47 (57.3%) | |
| Black not of Hispanic American Origin | 23 (5.6%) | 21 (7.6%) | 2 (3.6%) | | - | |
| Asian or Pacific Islander | 41 (10.0%) | 23 (8.4%) | 6 (10.9%) | | 12 (14.6%) | |
| Hispanic/Latino | 25 (6.1%) | 16 (5.8%) | 3 (5.5%) | | 6 (7.3%) | |
| Other | 30 (7.3%) | 19 (6.9%) | 3 (5.5%) | | 8 (9.8%) | |
| Histological Grade |  |  |  | |  | |
| Grade 1 | 1 (0.2%) | - | - | | 1 (1.2%) | |
| Grade 2 | 64 (15.5%) | - | 27 (49.1%) | | 37 (45.1%) | |
| Grade 3 | 44 (10.7%) | - | 27 (49.1%) | | 17 (20.7%) | |
| Grade 4 | 285 (69.2%) | 263 (95.6%) | - | | 22 (26.8%) | |
| Unknown | 21 (4.4%) | 12 (4.4%) | 1 (1.8%) | | 4 (4.9%) | |
| Molecular Markers |  |  |  | |  | |
| IDH Status |  |  |  | |  | |
| Mutated | 137 (33.5%) | 0 (0.0%) | 55 (100.0%) | | 82 (100.0%) | |
| Wildtype | 275 (66.5%) | 275 (100.0%) | 0 (0.0%) | | 0 (0.0%) | |
| 1p/19q Codeletion |  |  |  | |  | |
| Present | 55 (13.3%) | 0 (0.0%) | 55 (100.0%) | | 0 (0.0%) | |
| Absent | 357 (86.7%) | 275 (100.0%) | 0 (0.0%) | | 82 (100.0%) | |
| ATRX Status |  |  |  | |  | |
| Lost | 73 (17.7%) | 2 (0.7%) | 0 (0.0%) | | 71 (86.6%) | |
| Retained | 339 (82.3%) | 273 (99.3%) | 55 (100.0%) | | 11 (13.4%) | |
| Patch Statistics |  |  |  | |  | |
| Average Patches per Patient, median [IQR] | 2430 (1260-3744) | 2052 (1224-3276) | 3708 (2340-5148) | | 2610 (1143-4023) | |
| Average Tumor Patches per Patient, median [IQR] | 921 (429-1449) | 929 (445-1674) | 1166 (627-2028) | | 734 (327-1552) | |

### Figure S1: Attention Threshold Ablation

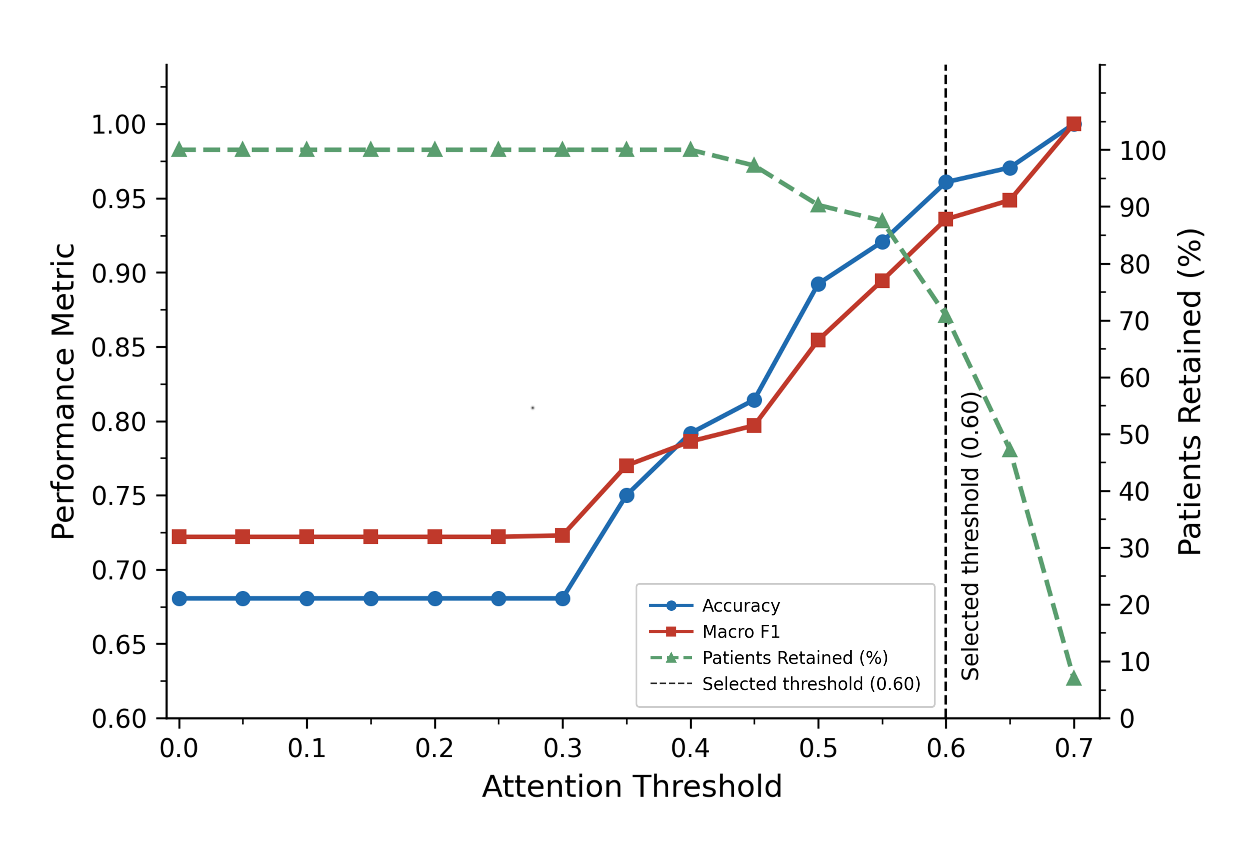

Results of an attention threshold ablation study demonstrating the relationship between patient accuracy, macro-F1 score, and retention rate versus attention threshold. The dashed vertical line denotes the selected attention threshold of 0.6.

### Figure S2: Molecular Probability Cutoffs and MLP Size Ablation

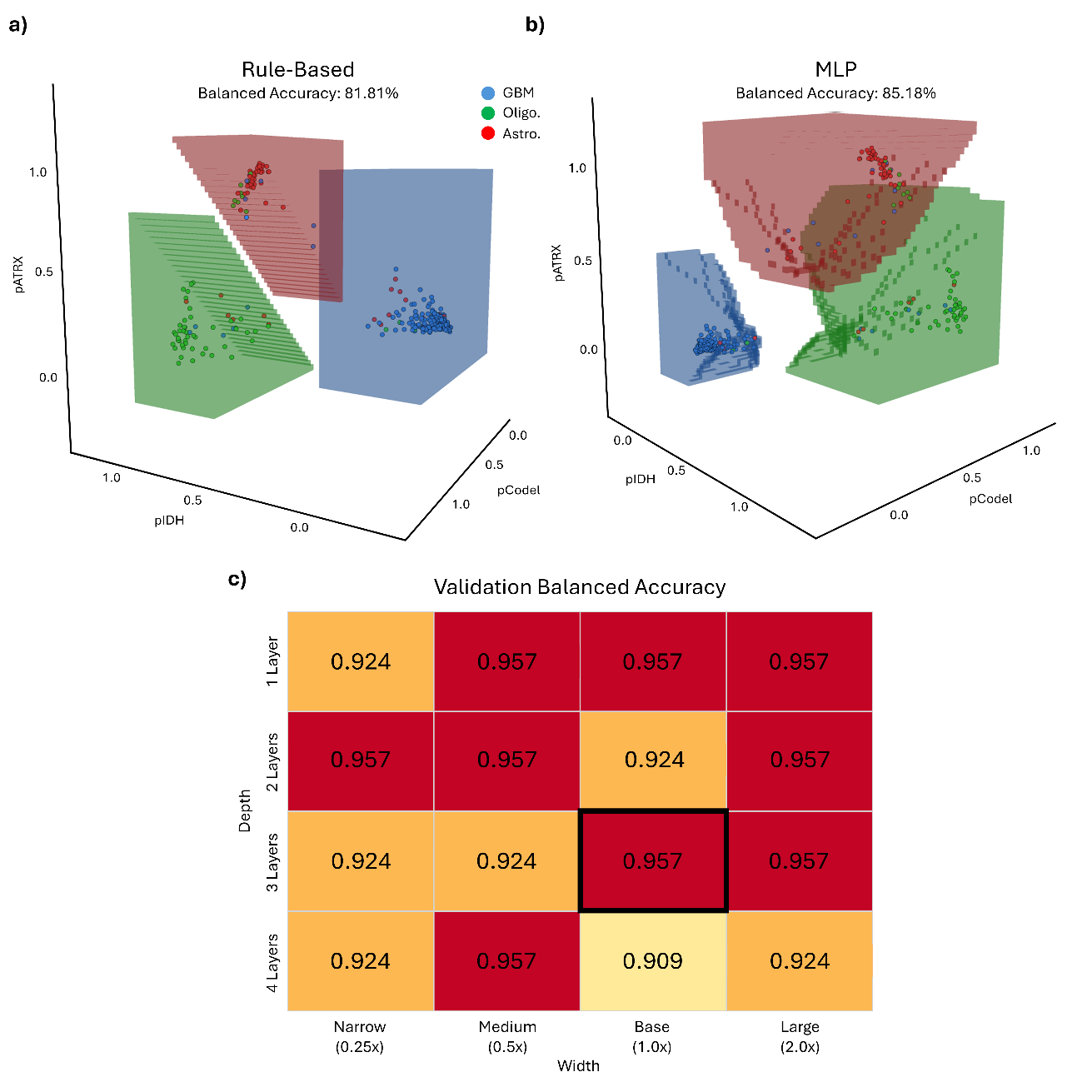

a) A 3-D visualization of the class cutoffs using the rule-based method seen in DeepGlioma, creating linear decision boundaries (Figure 1) b) A 3-D visualization of the class cutoffs determined by the final MLP trained on the three molecular probabilities output by the molecular MLP heads c) the results of an ablation study to determine the ideal size of the final MLP. The depth axis varies the number of layers, while the width axis varies the size of each layer relative to the base size. The base 3-layer model (highlighted in black) is 128-128-64.

### Figure S3: LIOCV Experimental Results

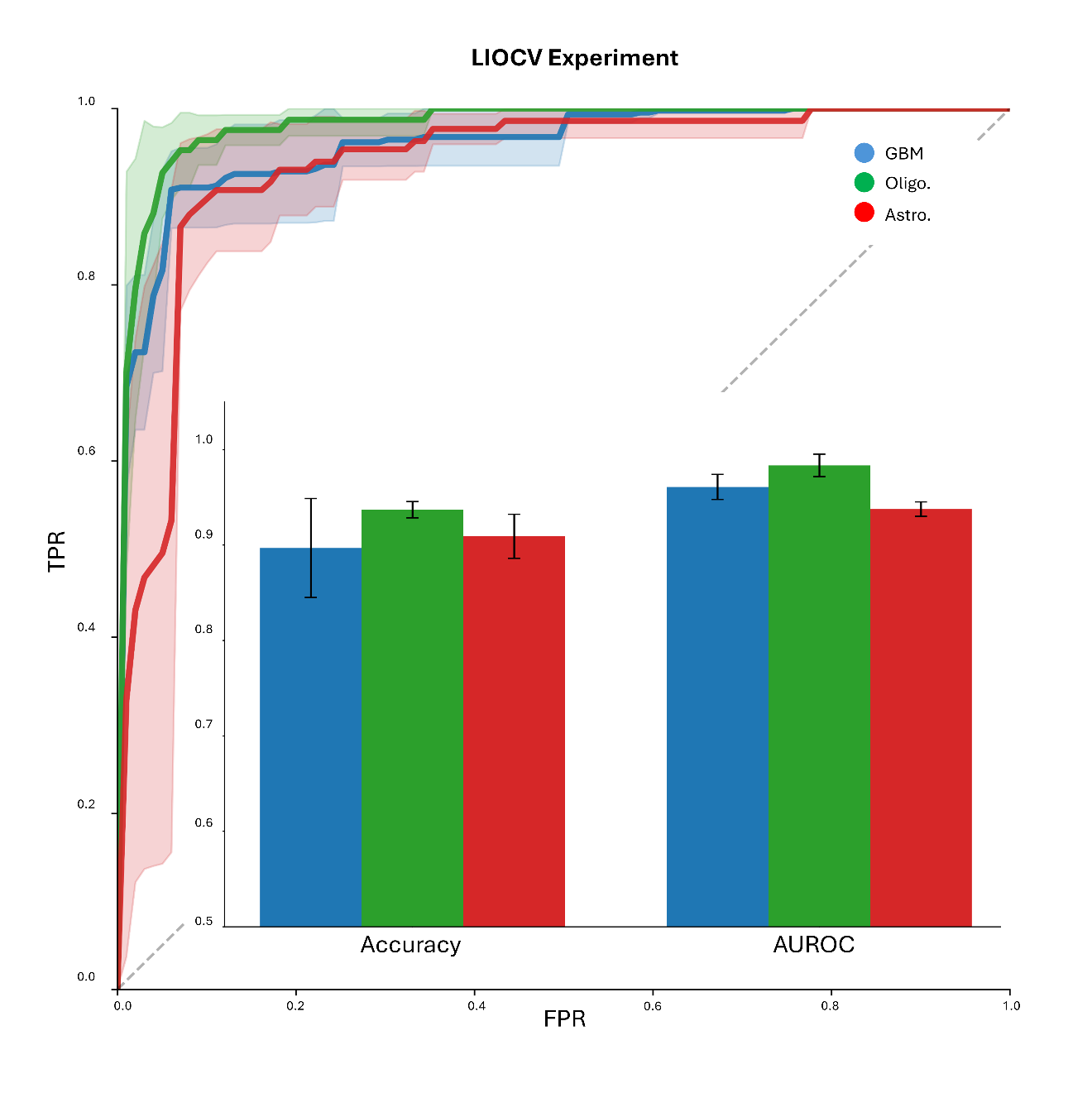

The results of a leave institution out cross validation (LIOCV) experiment on our test set, showing one-versus-rest area under the receiver operating curve (AUROC) and class accuracy with associated 95% confidence intervals. .

### Figure S4: Patch Count Versus Accuracy and Stability

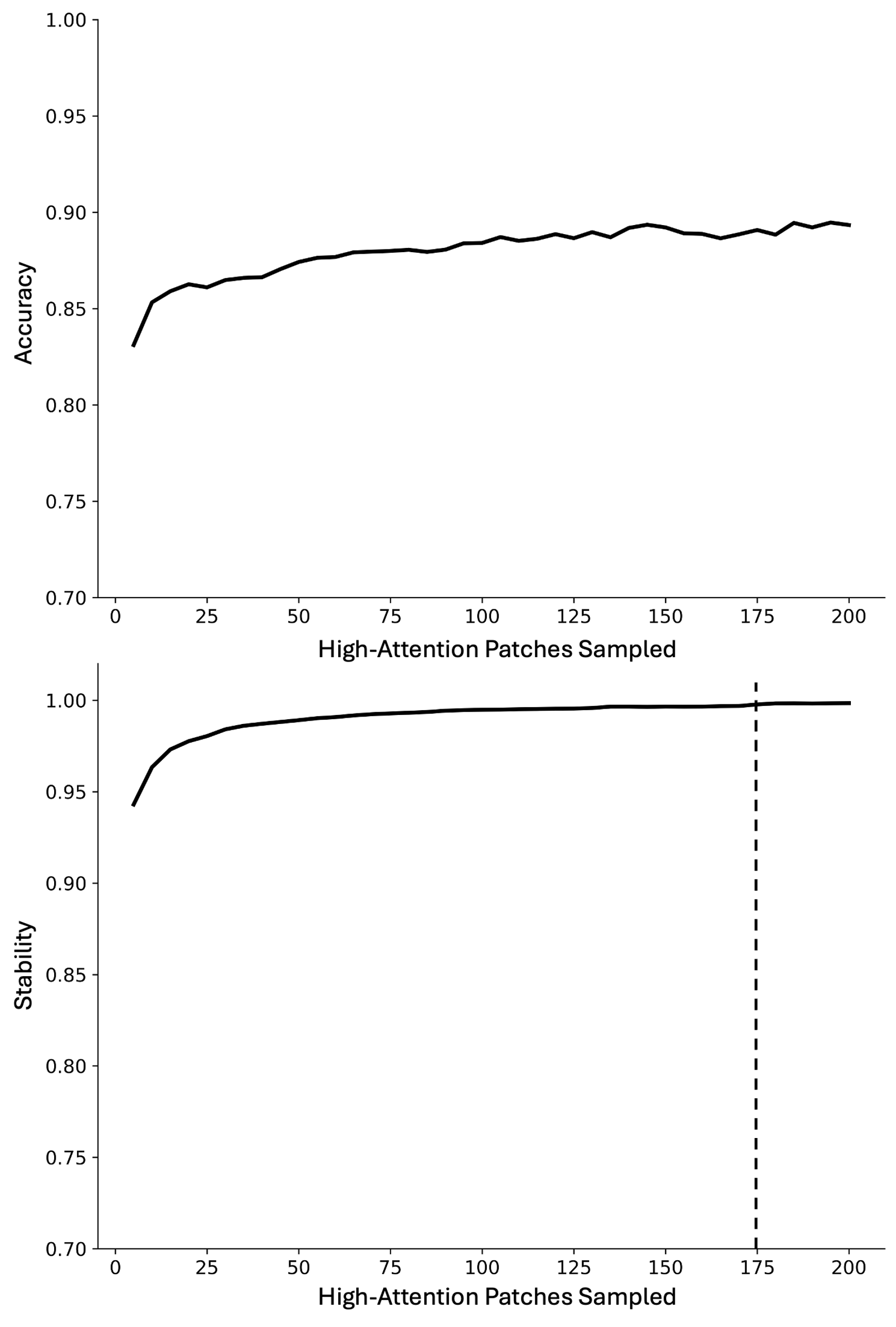

Graphs showing model accuracy (top) and prediction stability, or the percentage of patch samples from a patient that the model predicted as the same class as with all tissue from the same patient (bottom), versus the number of randomly sampled high-attention (attention > 0.6) patches. The vertical dashed line in the stability figure represents the cutoff for 99.5% stability (175 patches).

### Figure S5: Model Speed Experimental Results

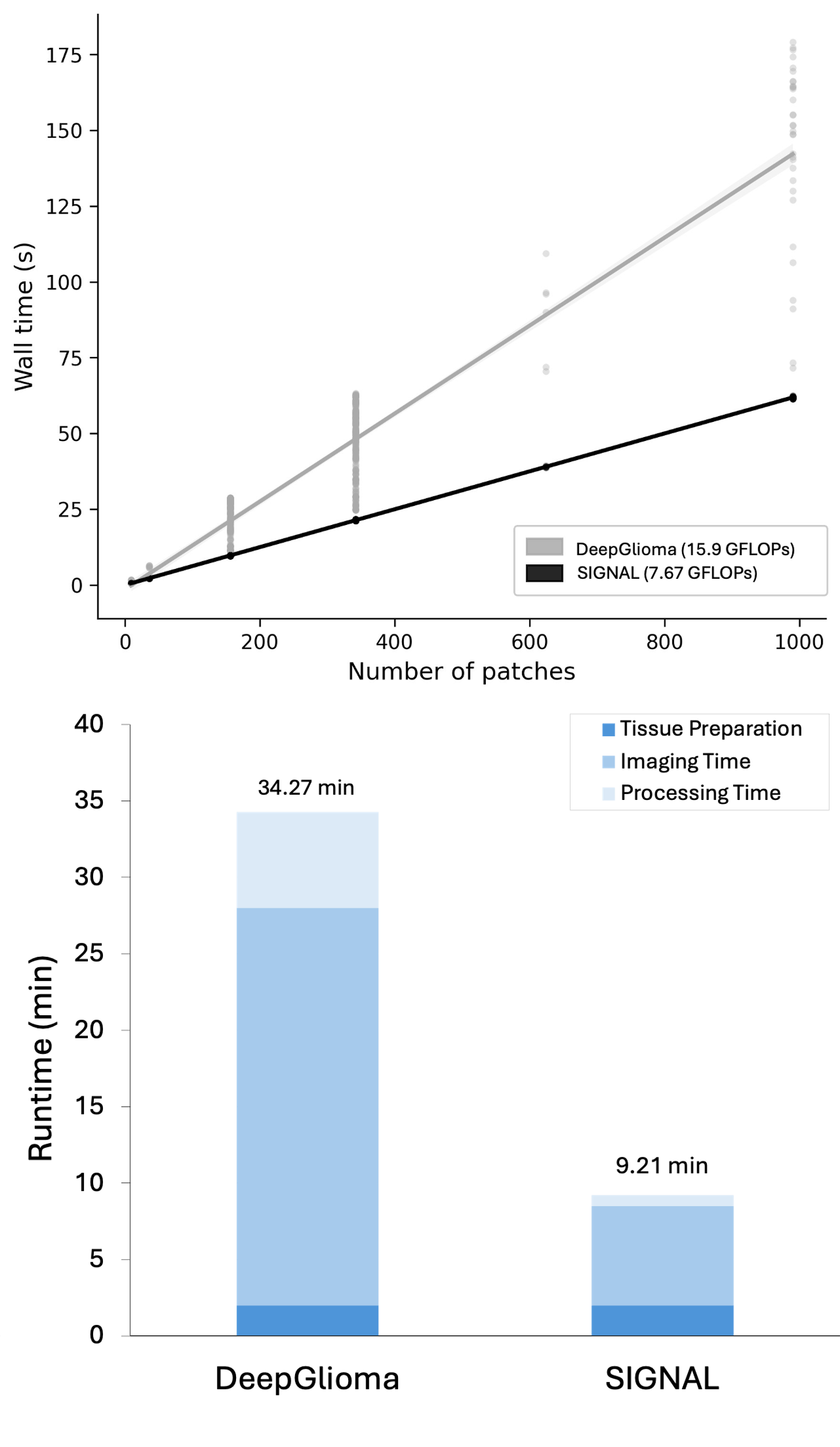

Whole-slide image processing speeds (top) for SIGNAL (black) and DeepGlioma (grey). A simulated comparison of intraoperative workflow times between DeepGlioma and SIGNAL (bottom). Based on the real-world workflow used for DeepGlioma, this simulation assumes DeepGlioma requires eight 2.5x2.8mm slides (mean number of slides acquired, 324 patches, 3.25 min/slide acquisition time) for prediction. SIGNAL has a 30% patch retention rate, meaning that on average, two 2.5x2.8mm slides are needed to reach the 175 patch 99.5% stability threshold.

### Figure S6: Bayesian Logistic Regression

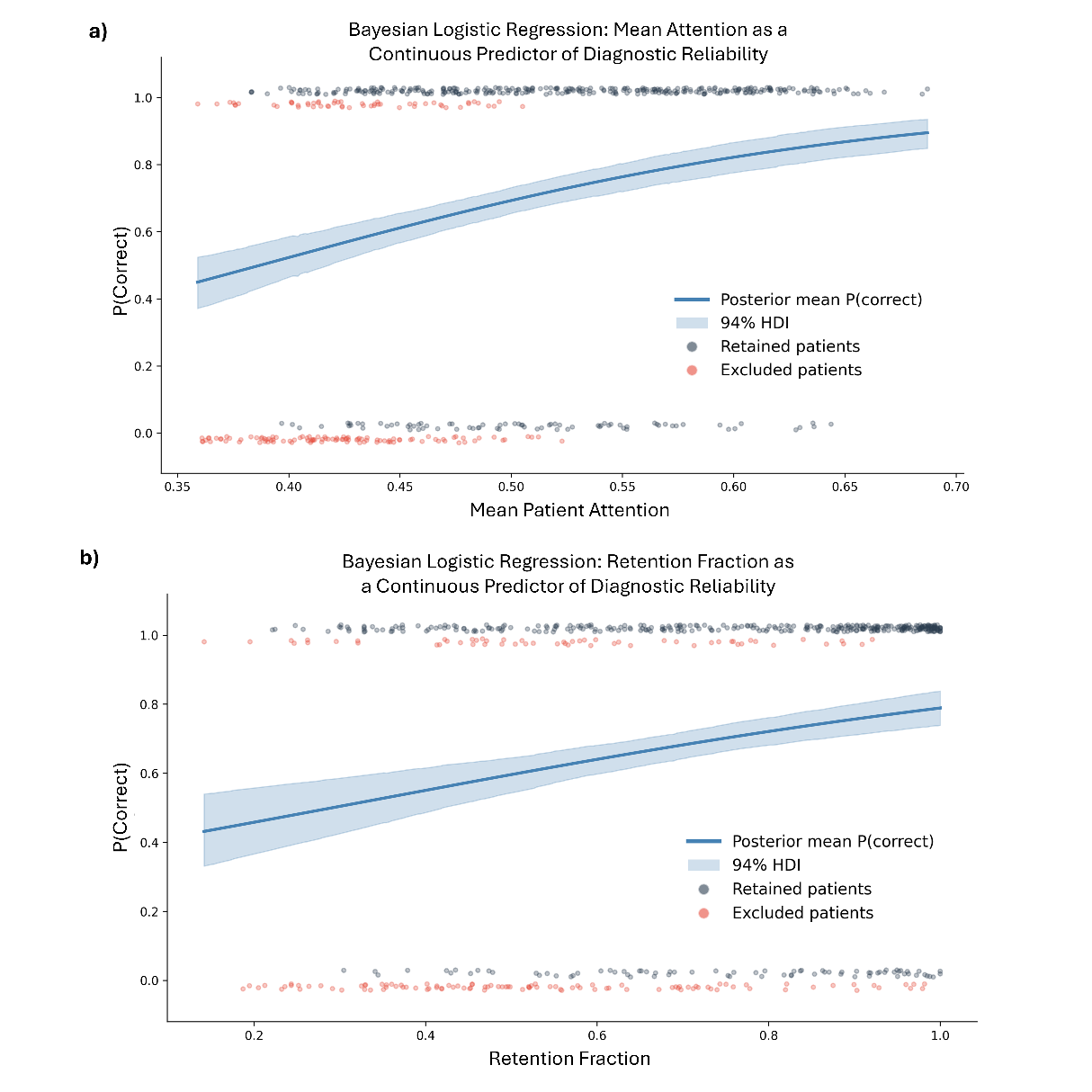

a) The Bayesian logistic regression curve for mean patient attention (prior to filtering) as a predictor of diagnostic accuracy, with the posterior probability mean probability curve and 94p% highest density interval (HDI) estimated via PyMC (4 chains x 2,000 draws, 1,000 tuning steps). Mean patient attention as strongly and credibly associated with correct classification (β = 7.22, 94% HDI [5.18, 9.34]; P(β > 0) = 1.0). b) The Bayesian logistic regression curve for patient retention fraction as a predictor of diagnostic accuracy, with the posterior probability mean probability curve and 94% highest density interval (HDI) estimated via PyMC. Patient retention fraction was also strongly and credibly associated with correct classification (β = 1.87, 94% HDI [1.11, 2.61]; P(β > 0) = 1.0).

### Figure S7: Normal Cadaveric Brain Tissue Sensitivity Analysis

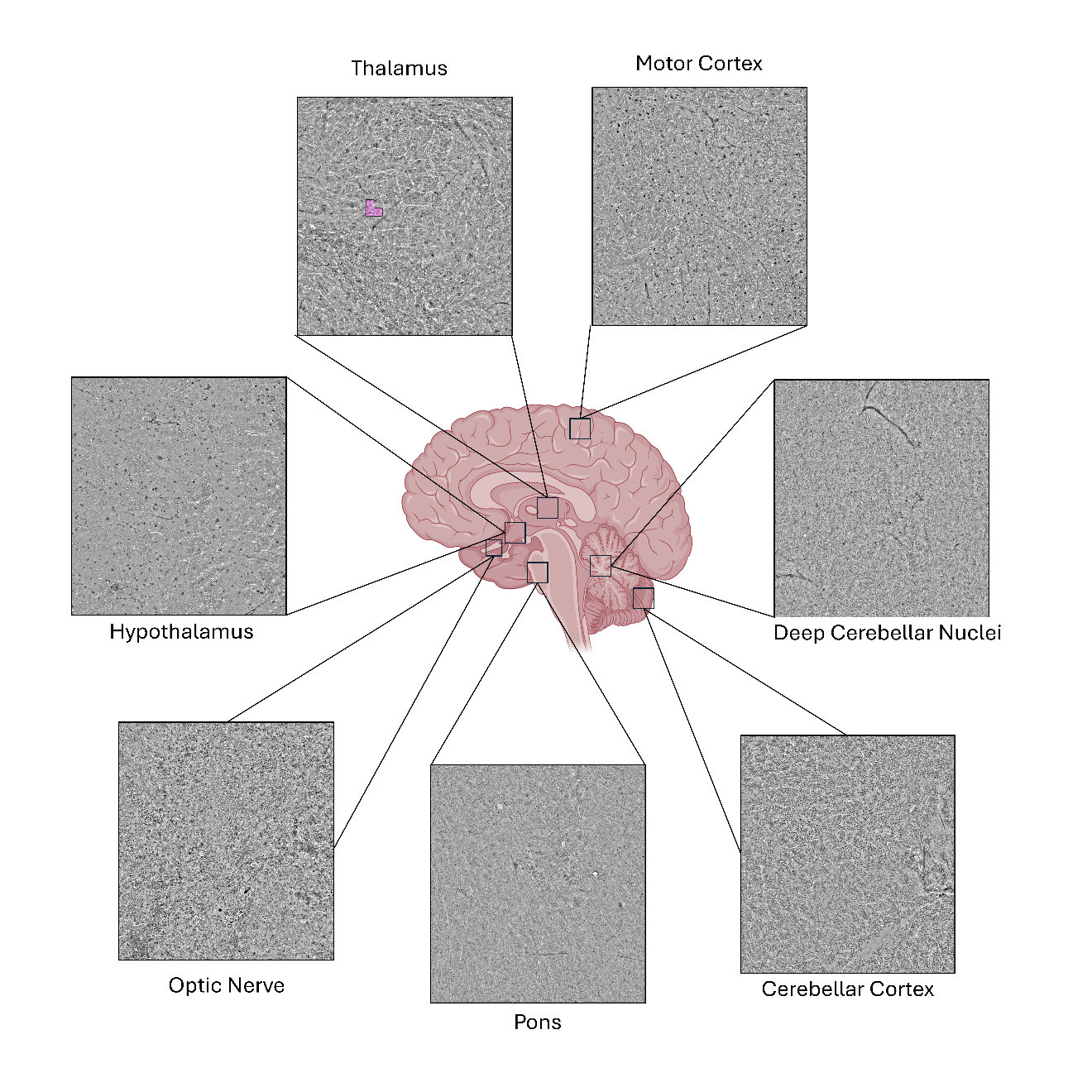

Example SRH images from normal cadaveric brain, colored by whether a patch meets the attention threshold (pink: attention > 0.6; grey: attention < 0.6). Of these images, only the thalamus contains patches retained by SIGNAL’s attention filter (0.13% by area), though it did not meet the 10% area coverage minimum for prediction.

### Figure S8: DeepGlioma Versus SIGNAL Filter Analysis

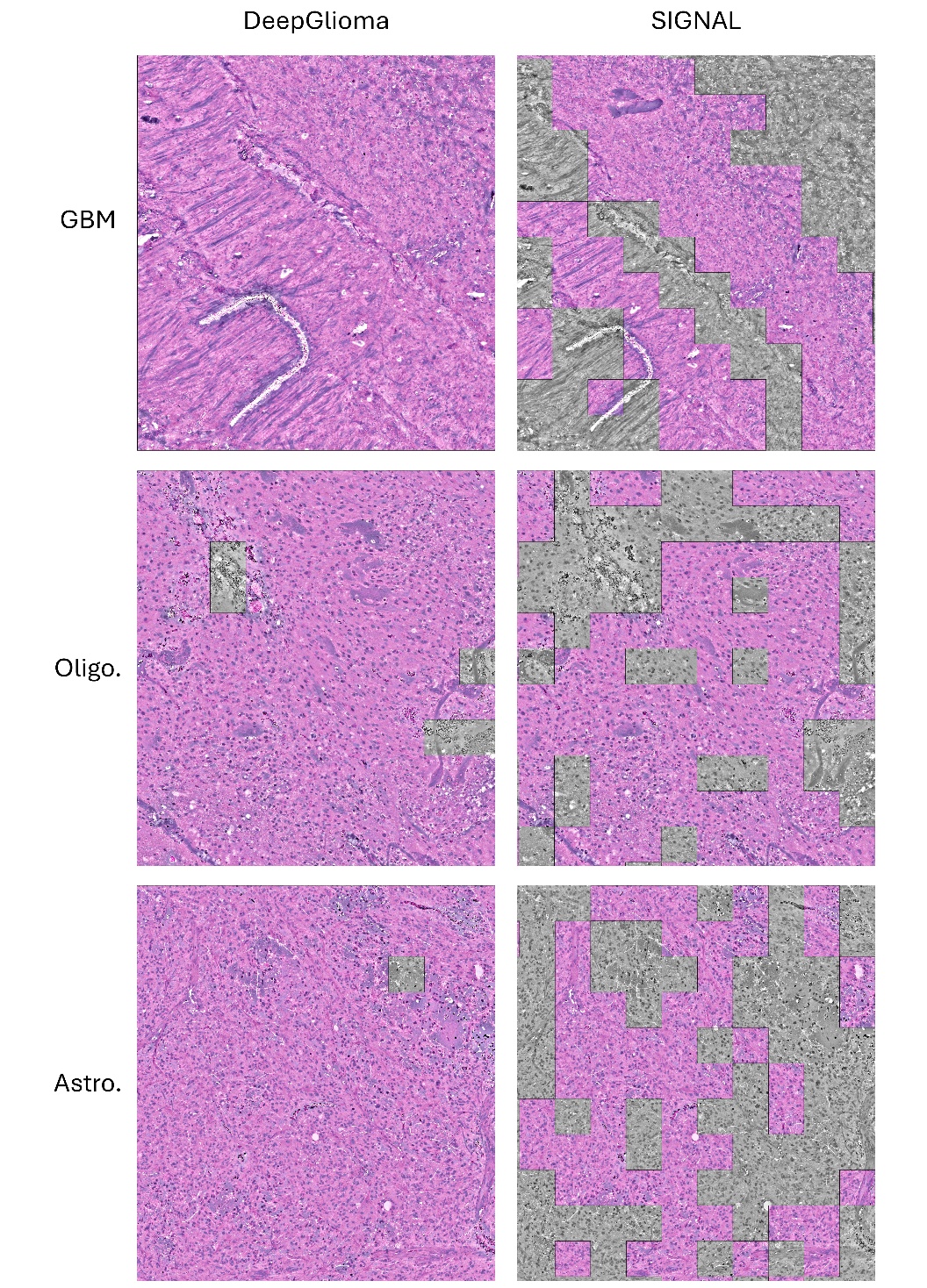

Examples of DeepGlioma and SIGNAL’s filtering mechanisms on different tumor subtypes. DeepGlioma’s tumor/non-tumor model shows much higher rates of tissue retention, including areas of higher blood coverage, fewer cells, and other features that lead to less favorable diagnostic conditions. SIGNAL’s attention mechanism, on the other hand, demonstrates a higher rate of filtering, with higher average retained tissue quality.

### MI-CLAIM Checklist

| **Before paper submission** | | | |
| --- | --- | --- | --- |
| **Study design (Part 1)** | **Completed:**  **page number** | | **Notes if not completed** |
| The clinical problem in which the model will be employed is clearly detailed in the paper. | ☑ | 3 |  |
| The research question is clearly stated. | ☑ | 3 |  |
| The characteristics of the cohorts (training and test sets) are detailed in the text. | ☑ | Suppl. Table 1 |  |
| The cohorts (training and test sets) are shown to be representative of real-world clinical settings. | ☑ | 4-5 |  |
| The state-of-the-art solution used as a baseline for comparison has been identified and detailed. | ☑ | 5 |  |
| **Data and optimization (Parts 2, 3)** | **Completed:**  **page number** | | **Notes if not completed** |
| The origin of the data is described and the original format is detailed in the paper. | ☑ | 3-4 |  |
| Transformations of the data before it is applied to the proposed model are described. | ☑ | Suppl. Methods |  |
| The independence between training and test sets has been proven in the paper. | ☑ | 3,5 |  |
| Details on the models that were evaluated and the code developed to select the best model are provided. | ☑ | Suppl. Methods |  |
| Is the input data type structured or unstructured? | ☐ Structured ☑ Unstructured | | |
| **Model performance (Part 4)** | **Completed:**  **page number** | | **Notes if not completed** |
| The primary metric selected to evaluate algorithm performance (eg: AUC, F-score, etc) including the justification for selection, has been clearly stated. | ☑ | 6 |  |
| The primary metric selected to evaluate the clinical utility of the model (eg PPV, NNT, etc) including the justification for selection, has been clearly stated. | ☑ | Suppl. Figure 6 |  |
| The performance comparison between baseline and proposed model is presented with the appropriate statistical significance. | ☑ | 6 |  |
| **Model Examination (Parts 5)** | **Completed:**  **page number** | | **Notes if not completed** |
| Examination Technique 1^a^: Saliency Maps (via Attention Correlations) | ☑ | 7 |  |
| Examination Technique 2^a^: Sensitivity Analyses (via multi-institutional external validation) | ☑ | 7 |  |
| A discussion of the relevance of the examination results with respect to model/algorithm performance is presented. | ☑ | 9 |  |
| A discussion of the feasibility and significance of model interpretability at the case level if examination methods are uninterpretable is presented. | ☑ | 6 |  |
| A discussion of the reliability and robustness of the model as the underlying data distribution shifts is included. | ☑ | 8-9 |  |
| *Common examination approaches based on study type:  * For studies involving exclusively structured data coefficients and sensitivity analysis are often appropriate  * For studies involving unstructured data in the domains of image analysis or NLP: saliency maps (or equivalents) and sensitivity analysis are often appropriate |  |  |  |
| **Reproducibility (Part 6): choose appropriate tier of transparency** | | | **Notes** |
| Tier 1: complete sharing of the code | | ☑ | Will be made publicly available on GitHub upon acceptance |
| Tier 2: allow a third party to evaluate the code for accuracy/fairness; share the results of this evaluation | | ☑ |  |
| Tier 3: release of a virtual machine (binary) for running the code on new data without sharing its details | | ☑ |  |
| Tier 4: no sharing | | ☑ |  |

PPV: Positive Predictive Value

NNT: Numbers Needed to Treat

^a^ Common examination approaches based on study type: for studies involving exclusively structured data, coefficients and sensitivity analysis are often appropriate; for studies involving unstructured data in the domains of image analysis or natural language processing, saliency maps (or equivalents) and sensitivity analyses are often appropriate. Select 2 from this list or chose an appropriate technique, document each technique used on the appropriate line above.
